# Anti-JOVI.1 antibody to detect clonal T cell populations: implementation into a diagnostic flow cytometry laboratory and correlation with clinical findings

**DOI:** 10.1101/2024.03.20.24304408

**Authors:** Benjamin Reardon, Jennifer Hsu, Sandy Smith, Riana van der Linde, David A. Brown, Elizabeth Tegg, Sarah C. Sasson

**Affiliations:** Department of Laboratory Haematology, Institute of Clinical Pathology and Medical Research (ICPMR), NSW Health Pathology, Westmead Hospital, Sydney, NSW, Australia; Flow Cytometry Unit, Institute of Clinical Pathology and Medical Research (ICPMR), NSW Health Pathology, Westmead Hospital, Sydney, NSW, Australia; Department of Immunopathology, Institute of Clinical Pathology and Medical Research (ICPMR), NSW Health Pathology, Westmead Hospital, Sydney, NSW, Australia; Westmead Clinical School, Sydney Medical School, University of Sydney, Australia; Centre for Immunology and Allergy Research, Westmead Institute for Medical Research, University of Sydney, Australia; University of Newcastle, University Drive Callaghan, NSW, Australia

## Abstract

The development of a high through-put flow cytometric assay for the identification of clonal T cells has proved challenging. We assessed the surface expression of a specific T Cell Receptor β-chain constant region using conjugated anti-JOV1.1 monoclonal antibodies to identify clonal T cell populations in a large diagnostic flow cytometry laboratory within a quaternary referral hospital. 37 cases were analysed. We identified 15 cases of clonal JOVI.1 expression, 7 of which had a consensus diagnosis of T-cell lymphoproliferative disease (TLPD). The remaining 22 cases had polyclonal JOVI.1 expression, none of which had a consensus diagnosis of TLPD, resulting in a sensitivity of 100% and specificity of 73%. When clonal NK-T cells were excluded, specificity further improves to 97%. These results provide real-world data and support the widespread adoption of this assay into diagnostic use.

## Introduction

T cell lymphoproliferative disease (TLPD) are uncommon and heterogenous in their clinical presentation and biological features^1^. Confirmation of clonal T cell expansion by flow cytometry can be challenging, as neoplastic and reactive T cell populations can have similar morphology and immunophenotype^2^. Combined with this, there is a lack of accessible and reproducible routine diagnostic tests available^2^. The advent of multiparameter flow cytometry is increasingly demonstrating its capacity to identify discrete T cell populations of uncertain significance. It may therefore have the capacity to determine whether these populations are monotypic as well as their clinical significance^3^.

Molecular assays to determine T cell clonality amplify specific regions of the T cell receptor (TCR), most often the αβ compared with γδ subsets. Similar to the kappa and lambda subtypes of the B cell receptor, the TCR β-chain constant region is encoded by either TRBC1 or TRBC2^2^. The use of a monoclonal antibodies (mAbs) directed towards TRBC1 and TRBC2, denoted as JOVI.1 and JOVI.2 respectively, can identify monotypic αβ T cell populations^4^. Anti-JOVI.1 mAbs have been used to identify Sezary syndrome, where the need for additional molecular T cell clonality testing has been negated^3^. However, TCR monotypy may also identify oligoclonal expansion of T-cell populations^5^.

In this study we assessed the utility of anti-JOVI.1 surface labelling to determine clonality of suspicious or aberrant T cell populations in a routine diagnostic flow cytometry laboratory and correlated these findings with available concurrent molecular and histopathological analysis.

## Method

We studied consecutive adult cases where T cell immunophenotyping by flow cytometry was performed from November 2021 to February 2022. All samples from patients over 18 years who had abnormalities relating to any of the following were further analysed: CD4:CD8 ratio <0.8 or >2.0, absolute CD4^+^ or CD8^+^ T cell count >3.0×10^9^/L, a history of TLPD, eosinophilia >1.0×10^9^/L, T cell population of interest >1.0×10^9^/L on blood or bone marrow sample or at clinician request were included. All samples from patients under 18 years of age were excluded. Samples were analysed on a Gallios flow cytometer (Beckman Coulter, California, USA) using three 10-colour panels (see Supplementary Tables S1A, S1B, and S1C). Further interrogation of αβ and γδ TCR was also performed in select cases (see Supplementary Table S1D). A minimum of 100,000 CD45^+^ events were required. Instrument quality control checks were performed daily, with compensation calibration performed weekly. NK-T cells were defined as co-expressing CD3 and CD56, with or without CD16 expression. JOVI.1 staining and analysis was performed by a scientist with specific expertise in flow cytometry who was blinded to the consensus diagnosis. Monotypic T cell populations were defined by JOVI.1 expression being either ≥90% or ≤10% positive on T cells, which are more stringent than previously published thresholds of ≥85% or ≤15% ^6^.

A corresponding clinical dataset was collected including age, gender, full blood count and differential, lymphocyte count, lymphocyte subsets, histopathological diagnosis, clinical and treatment history. The consensus diagnosis of TLPD was determined by the treating clinician using all available results at the time of at the time of flow cytometric analysis. Descriptive statistics were used to analyse the demographic data. Differences in total lymphocyte counts between groups was determined by a non-parametric Mann-Whitney test with a p<0.05 considered statistically significant. All statistical analysis was performed using GraphPad software (Prism; Boston, USA). Ethics approval was obtained as per local institutional policy.

## Results

Thirty-seven samples were analysed, derived from: peripheral blood (N=27), bone marrow (N=7), pleural fluid (N=1) and other biopsy sites (N=2). The median age of patients was 65 years. There were 18 male and 19 female patients. The median total peripheral blood lymphocyte count was 1.7×10^9^/L, and median CD4:CD8 ratio was 0.9. Consensus diagnosis were comprised of TLPD (N=7; Including four cases of Sezary Syndrome, two of T cell Large Granular Lymphocytic Leukaemias and one of T cell Prolymphocytic Leukaemia), TLPD at a distant site without active disease within the sample sent for flow cytometric analysis (e.g., mycosis fungoides in skin but not in peripheral blood or bone marrow. N=6), B-cell lymphoma (N=6), immune mediated cytopenias (N=5), solid organ malignancy (N=3), psychotic illness (N=2), infection (N=2),, dermatitis (N=1), lymphocytosis of unclear aetiology (N=1), multiple myeloma (N=1), diagnosis unknown (N=2), data missing (N=1). Supplementary table S2 summaries all cases and their consensus diagnoses.

Monotypic JOVI.1 expression was detected in 8 of the 37 cases. 7 of the 8 cases had a consensus diagnosis of TLPD. An example of a positive result is shown in Figure 1. Polytypic JOVI.1 expression was detected in the remaining 29 cases, all of which did not have a diagnosis of TLPD, resulting in an overall test sensitivity of 100% and specificity of 97% (Table 1b).

**Table 1:** Sensitivity and Specificity of JOVI.1 expression for the detection of T cell Lymphoproliferative Disease (TLPD)

**a) All samples**
|  | Consensus Diagnosis: TLPD<br>(N=7) | Consensus diagnosis: Non-TLPD<br>(N=30) |
| --- | --- | --- |
| JOVI.1 Clonal | 7 | 8 |
| JOVI.1 Polyclonal | 0 | 22 |

**b) Excluding small secondary NK-T cell populations**
|  | Consensus Diagnosis : TLPD<br>(N=7) | Consensus diagnosis: Non-TLPD<br>(N=30) |
| --- | --- | --- |
| JOVI.1 Clonal | 7 | 1 |
| JOVI.1 Polyclonal | 0 | 29 |

*Sensitivity=100%; Specificity= 97%*

**Figure 1:**
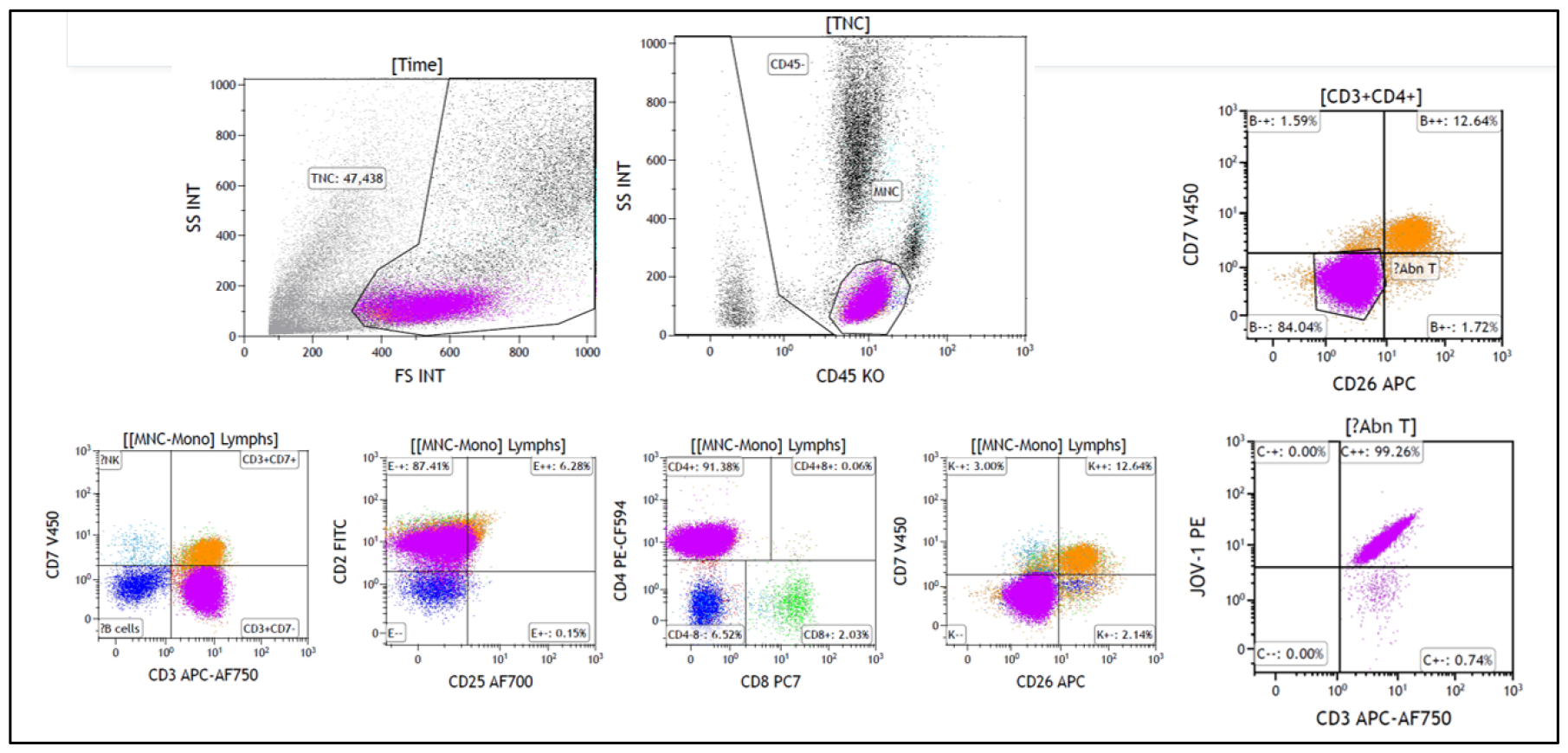
Anti-JOVI.1 monoclonal antibody staining for detection of monotypic T-cell population. Flow cytometric histogram showing an example of a monotypic T cell population (purple) expressing CD45 with low side scatter (SS) in the lymphocyte region. This population expresses CD3 and CD4 and lacked CD7 and CD26. 99% of this population expresses JOVI.1. Findings are consistent with a diagnosis of Sezary syndrome.

A secondary finding was that NK-T cell populations with monotypic JOVI.1 expression were found in seven cases, none of which had an aberrant immunophenotype. These were found in the setting of a range of malignant (N=3) and non-malignant (N=4) conditions. Of patients with NK-T cells monotypic for JOVI.1 in the setting of a malignancy, all patients were on disease-specific treatment. If monotypic NK-T cells were included in the analysis, specificity of the JOVI.1 antibody is reduced to 73% (Table 1a).

Prominent γδ T-cell populations were evident in three patients. By definition, γδ T-cells do not express the TCR β-chain constant region and thus JOVI.1 monoclonal antibodies will not bind this TCR. None of these three patients had a consensus diagnosis of TLPD.

There were no differences in absolute total lymphocyte count in those with TLPD compared with those without (p = 0.10, Fig. 2). TCR gene rearrangement studies were available in two cases for further correlation. This included one case of a positive TCR gene rearrangement study in a patient with T cell large granular lymphocyte leukaemia, where monotypic JOVI.1 expression was also demonstrated, and the second in a patient with chronic B cell lymphocytic leukaemia with a negative TCR rearrangement study and polytypic JOVI.1 expression.

**Figure 2:**
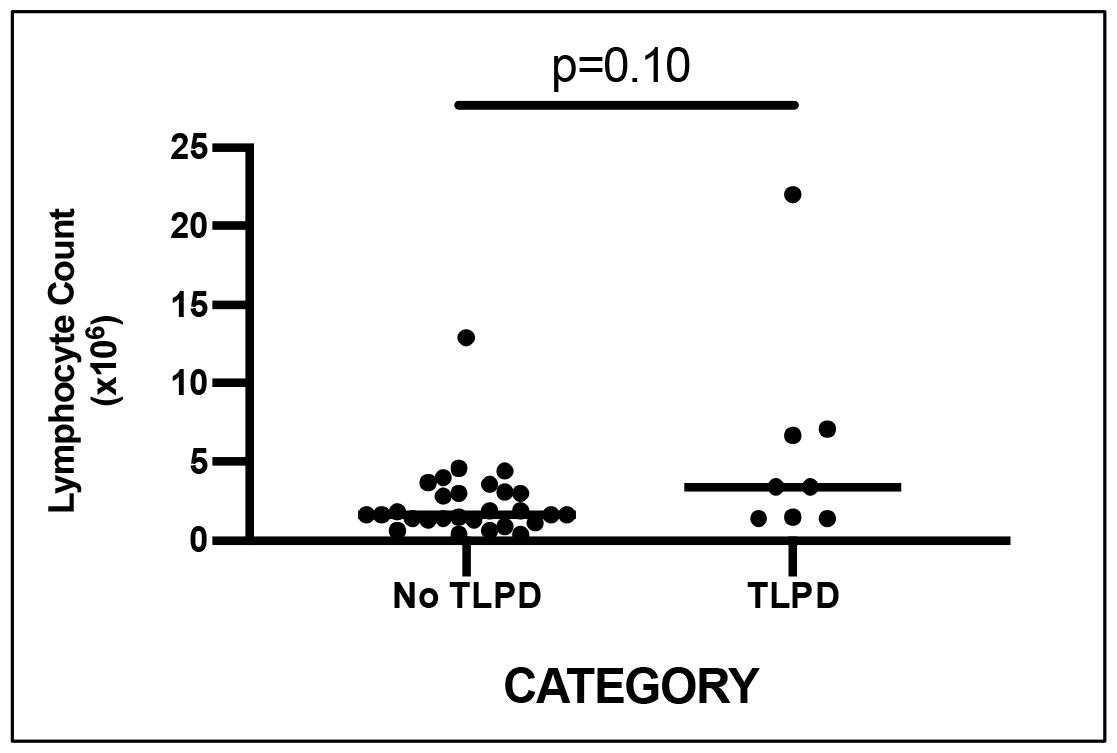
Total lymphocyte count and presence of T cell lymphoproliferative disease.

## Discussion

This study provides important ‘real-world’ data regarding the implementation of JOVI.1 testing into a large diagnostic laboratory, including analysis of a variety of sample types from patients with a broad range of underlying conditions.

Overall, these results demonstrate the use of anti-JOVI.1 mAbs have a high sensitivity and specificity for the detection of TLPD. In the single case where monotypic JOVI.1 expression could not be correlated to a consensus diagnosis of TLPD, documentation relating to the histopathological and clinical diagnosis was unable to be obtained. Our sensitivity of 100% is higher than previous reports and likely reflects our criteria for selecting cases for JOVI.1 testing, rather than testing as part of a screening algorithm^5-7^.

This work highlights two important factors, that being the need to identify and exclude from analysis both γδ- and NK-T cell populations prior to JOVI.1 analysis on αβ T cells. By definition, γδ will not bind JOVI.1 mAb, and their inclusion has the potential to lead to false positive reports of T cell populations that are <10% positive for JOVI.1 expression.

Additionally, this work identified that small *secondary* subpopulations of NK-T cells can demonstrate monotypic JOVI.1 expression. Seven cases of monotypic JOVI.1 NK-T cells were found, two of which expressed CD4, and three of which expressed CD8. All of these populations had an absolute lymphocyte count <1.0×10^9^/L. These cases are consistent with T cell clonopathy of undetermined significance (TCUS), and were found in patients with co-morbid malignancy on treatment (non-TLPD) and non-malignant conditions including infection and psychotic illness. This work provides evidence that small clonal NK-T cell populations occur in the setting of anti-tumour and anti-infective immunity or associated with treatment^8, 9^. Identification and exclusion of the NK-T cell compartment prior to JOVI.1 analysis improves the specificity of the assay (Table 1). Our work has provided independent verification of the clinical utility of JOVI.1 testing in a diagnostic environment, and has provided important methodological considerations regarding the need to exclude γδ- and NK-T cells prior to analysis. However, we acknowledge the limitations of this work being a single-centre study with a relatively small sample size.

Until recently, the main methods to detect TCR clonality have been restriction enzyme nuclease digestion of diagnostic tissue followed by gel electrophoresis and Southern blotting. Although cumbersome, this technique is still widely accepted as the gold standard for T cell clonality determination. Other molecular methods include polymer chain rection (PCR)-based techniques and next generation sequencing. Compared to these techniques, flow cytometry offers a faster turnaround time, however the previous TCR Vβ assay, which involved staining suspicious populations with mAbs that covered ∼65% of TCR Vβ was both labour intensive and expensive. The recent development of the anti-JOVI.1 mAb, applied in multiparameter flow cytometry reduces the labour and expense associated with the TCR Vβ assay and has a fast turnaround time with a reasonably high sensitivity and specificity for a variety of diagnostic tissues.

## Conclusion

Our work combined with that of other groups^3-7^ provides compelling evidence for more widespread adoption of this assay into diagnostic use.

## Data Availability

All data produced in the present work are contained in the manuscript.

## Notes

### Competing Interest Statement

The authors have declared no competing interest.

### Funding Statement

This study did not receive any funding

### Author Declarations

Ethics approval was obtained from Western Sydney Local Health District Research Office on 09/02/2022. The committee granted a waiver for the usual requirement of consent for the use of re-identifiable information by New South Wales (NSW) agencies, in line with the State Privacy Commissioner's Guidelines for Research and the Health Records and Information Privacy Act 2022 (NSW).

