## Supplementary material for "Anti-JOVI.1 antibody to detect clonal T cell populations: implementation into a diagnostic flow cytometry laboratory and correlation with clinical findings": Table S1

**Table S1A: T cell Panel 1**

| **Antibody** | **Colour** | **Clone** |
| --- | --- | --- |
| CD2 | FITC | RPA-2.10 |
| JOVI-1 | PE | JOVI.1 |
| CD4 | PE-CF594 | RPA-T4 |
| CD5 | PerCP | UCHT2 |
| CD8 | PC7 | SK1 |
| CD26 | APC | M-A261 |
| CD25 | AF700 | M-A251 |
| CD3 | APC AF750 | UCHT1 |
| CD7 | V450 | M-T701 |
| CD45 | KO | J33 |

**Table S1B: T cell Panel 2**

| **Antibody** | **Colour** | **Clone** |
| --- | --- | --- |
| CD57 | FITC | NK-1 |
| JOVI-1 | PE | JOVI.1 |
| CD4 | PE-CF594 | RPA-T4 |
| CD5 | PerCP | UCHT2 |
| CD8 | PC7 | SK1 |
| CD94 | APC | HP-3D9 |
| CD56 | AF700 | B159 |
| CD3 | APC AF750 | UCHT1 |
| CD16 | PB | 3G8 |
| CD45 | KO | J33 |

**Table S1C: T cell Panel 3**

| **Antibody** | **Colour** | **Chrome** |
| --- | --- | --- |
| CD2 | FITC | RPA-2.10 |
| CD7 | PE | M-T701 |
| CD4 | PE-CF594 | RPA-T4 |
| CD5 | PC5.5 | BL1a |
| CD19 | PC7 | J3-119 |
| CD56 | APC | B159 |
| CD3 | AF700 | UCHT1 |
| CD8 | APC Cy7 | SK1 |
| CD16 | PB | 3G8 |
| CD45 | KO | J33 |

**Table S1D: T cell Panel 4**

| **Antibody** | **Colour** | **Clone** |
| --- | --- | --- |
| ab | FITC | WT31 |
| gd | PE | 11F2 |
| CD4 | PE-CF594 | RPA-T4 |
| CD5 | PC5.5 | BL1a |
| CD8 | PC7 | SK1 |
| CD3 | APC | UCHT1 |
| CD7 | V450 | M-T701 |
| CD45 | KO | J33 |
