## Supplementary material for "Anti-JOVI.1 antibody to detect clonal T cell populations: implementation into a diagnostic flow cytometry laboratory and correlation with clinical findings": Table S2

**Table S2: Summary of cases and consensus diagnosis**

| **No.** | **Tissue Source** | **Age Range (years)** | **Gender** | **Lympho-cyte count**  **(x10^9/L)** | **CD4: CD8 Ratio** | **Diagnosis** |
| --- | --- | --- | --- | --- | --- | --- |
| 1 | PFluid | 66-70 | F | 0.4 | 0.36 | Metastatic melanoma on immunotherapy |
| 2 | PB | 56-60 | F | 3 | 0.64 | Lymphocytosis of unclear cause |
| 3 | BM | 60-65 | M | 1.4 | 4.4 | Cirrhosis |
| 4 | PB | 46-50 | F | 1.6 | 0.33 | Mycobacterium avium complex pulmonary infection with lymphadenopathy |
| 5 | PB | 51-55 | M | 12.9 | 0.07 | Peripheral T cell lymphoma, NOS |
| 6 | PB | 16-20 | F | 1.9 | 1.26 | Psychotic illness |
| 7 | PB | 66-70 | F | 1.4 | 1.38 | T cell prolymphocytic leukaemia |
| 8 | BM | 41-45 | M | 4 | 0.48 | Hairy Cell Leukaemia |
| 9 | PB | 16-20 | F | 1.5 | 1.7 | Psychotic illness |
| 10 | PB | 51-55 | M | 4.4 | 2.22 | Eczema |
| 11 | PB | 71-75 | M | 1.1 | 0.57 | Chronic lymphocytic leukaemia |
| 12 | PB | 71-75 | F | 0.9 | 0.9 | Autoimmune neutropenia |
| 13 | PB | 61-65 | M | 3.7 | 1.41 | Lung adenocarcinoma |
| 14 | PB | 61-65 | M | 3.4 | 0.24 | Sezary Syndrome |
| 15 | PB | 61-65 | F | 1.5 | 1.86 | Para-aortic lymphadenopathy with fibroinflammation |
| 16 | PB | 66-70 | F | 1.3 | 5.8 | Sezary syndrome |
| 17 | PB | 71-75 | M | 1.6 | 2.14 | Diffuse large B cell lymphoma |
| 18 | Bx | 51-55 | F | **Data missing* | 15 | Reactive lymphoid hyperplasia |
| 19 | PB | 76-80 | F | 1.4 | 2.69 | Sezary Syndrome |
| 20 | PB | 51-55 | M | 3 | 0.57 | Autoimmune cytopenias |
| 21 | BM | 46-50 | M | 0.6 | 0.74 | Peripheral T cell lymphoma, NOS |
| 22 | PB | 76-80 | F | 1.6 | 3.4 | **Data missing* |
| 23 | PB | 71-75 | F | 4.6 | 0.37 | Dermatitis |
| 24 | BM | 76-80 | M | 0.6 | 0.54 | Peripheral T cell lymphoma, NOS |
| 25 | PB | 81-85 | F | 6.7 | 39.15 | Sezary Syndrome |
| 26 | PB | 66-70 | F | 1.9 | 0.5 | Autoimmune neutropenia |
| 27 | PB | 71-75 | M | 1.4 | 1.7 | Mycosis fungoides |
| 28 | PB | 71-75 | F | 1.8 | 0.37 | Diffuse large B cell lymphoma |
| 29 | Bx | 26-30 | M | 0.4 | 9.1 | EBV positive Hodgkin’s lymphoma |
| 30 | PB | 61-65 | M | 3.1 | 1.26 | Metastatic renal cell carcinoma |
| 31 | PB | 51-55 | F | 3.6 | 2.39 | Mycosis fungoides |
| 32 | BM | 76-80 | M | 22 | 0.07 | T cell granular lymphocyte leukaemia |
| 33 | BM | 76-80 | M | 2.8 | 0.18 | Macrocytic anaemia |
| 34 | BM | 66-70 | M | 7.1 | 0.18 | T cell granular lymphocyte leukaemia |
| 35 | PB | 71-75 | F | 1.3 | 0.55 | IgG kappa multiple myeloma |
| 36 | PB | 41-45 | M | 1.6 | 0.99 | Cutaneous B cell lymphoma |
| 37 | PB | 61-65 | M | 3.4 | 0.34 | Relapsed Sezary Syndrome post allogeneic stem cell transplant |

Key: M male, F female, PB peripheral blood, BM bone marrow, Bx lymph node biopsy, PFluid pleural fluid, EBV Ebstein Barr virus, NOS not otherwise specified
